# HIV, Nephrotoxic Medications, and Chronic Kidney Disease: Prevalence, Risk Factors, and Mediation Analyses Among People With and Without HIV Enrolled in the Multicenter AIDS Cohort Study (MACS) / Women’s Interagency HIV Study (WIHS) Combined Cohort Study

**DOI:** 10.1101/2025.10.30.25339018

**Authors:** Yue Pan, Dominique Musselman, Zain Mithani, Weiqun Tong, Joseph Margolick, Frank J. Palella, Matthew J. Mimiaga, Kaitlin Bodnar, Deborah Konkle-Parker, Gina Wingood, Daniel Westreich, Eric Seaberg, Signe Lauren, Mardge Cohen, Michelle M. Estrella, Amanda Spence, Tracey Wilson, Michael Ross, Dan Feaster, Maria L. Alcaide, Deborah L. Jones

## Abstract

**Background:** Chronic kidney disease (CKD) affects over 37 million adults in the United States, and people living with HIV (PLWH) are at greater risk for progression to end-stage kidney disease. Although both conditions are common among PLWH, the potential pathways through which depression and use of medications with nephrotoxic potential may influence CKD development remain underexplored. We evaluated the relationships of depression and nephrotoxic medication use with CKD prevalence among PLWH, and investigated the potential mediating effects of these factors on the pathway to CKD among PLWH.

**Methods:** We analyzed longitudinal data from the Multicenter AIDS Cohort Study (MACS)/Women’s Interagency HIV Study (WIHS) Combined Cohort Study (MWCCS), collected between October 2018 and September 2021, to assess the prevalence of kidney dysfunction (eGFR <60 mL/min/1.73 m²) and its association with HIV serostatus. Generalized Estimating Equations (GEE) with a Poisson distribution and log link were used to estimate relative risks (RR) for CKD associated with HIV, depression, and other covariates. Counterfactual-based causal mediation analysis was conducted to assess whether depressive symptoms or nephrotoxic medication use partially explained the observed association between HIV and CKD.

**Results:** Among 2,530 participants [1,622 PLWH and 908 people living without HIV (PLWoH)], CKD prevalence was higher in PLWH (18.1%) compared to PLWoH (9.7%). GEE analysis revealed that HIV serostatus was significantly associated with an increased risk of CKD (RR = 1.37, 95% CI: 1.28-1.48, p < 0.0001). Other significant factors included age (RR = 1.03, 95% CI: 1.03-1.03, p < 0.0001), non-Hispanic Black (RR = 1.19, 95% CI: 1.11-1.27, p < 0.0001) compared to non-Hispanic White, diabetes (RR = 1.26, 95% CI: 1.17-1.35, p < 0.0001), and higher income (RR = 0.99, 95% CI: 0.98-1.00, p = 0.005). Mediation analysis indicated that nephrotoxic medication use, accounted for a small but significant proportion of the HIV- CKD association (indirect effect OR = 1.02, 95% CI: 1.00-1.03, p = 0.02).

**Conclusions:** While it is well established that PLWH have a higher prevalence of CKD compared to PLWoH, our findings suggest that nephrotoxic medication use may modestly amplify this risk. Although most of the risk appears to be attributable to the direct effects of HIV, these medications represent a modifiable contributor. PLWH receiving such treatments may benefit from closer kidney function monitoring. Future research should investigate additional psychosocial and behavioral contributors to CKD among PLWH, including depression.

## Introduction

People living with HIV (PLWH) enjoy markedly extended survival due to the routine use of virally suppressive antiretroviral therapy. However, PLWH remain at elevated risk for kidney-related complications, including a higher incidence of acute kidney injury and a greater burden of both general (e.g., hypertension and diabetes) and HIV-specific risk factors for chronic kidney disease (CKD). The prevalence of HIV-associated chronic kidney disease (CKD) varies widely by region, patient characteristics, reporting methods, and CKD definition. In North America and Europe, using a definition of eGFR <60 mL/min/1.73 m² or proteinuria, prevalence may reach 15% or higher [1].

The development of CKD in PLWH is often multifactorial. It may result from HIV-related immune dysfunction (e.g., prolonged immunosuppression, high viral load), genetic predisposition, and the use of certain antiretroviral medications [2]. Compared with the general population, PLWH experience a 2- to 20-fold greater risk of developing end-stage kidney disease [1, 3], with a disproportionately higher risk borne by Black individuals [4]. Even more fundamentally, HIV is directly kidney-tropic: in the pre-ART era, HIV-associated nephropathy (HIVAN), characterized by rapid progression to renal failure and collapsing focal segmental glomerulosclerosis was a leading cause of death among Black PLWH [5]. The kidney, as one of HIV’s primary targets, remains a critical focus for renal prevention and therapeutic strategies.

Depression is also common amongst PLWH, occurring in about 28% of PLWH who are older than 50 years of age [6]. Two recent community-based studies highlighted the relationship between depression and CKD. The China Health and Retirement Longitudinal Study enrolled 4763 participants with eGFR≥60 ml/min per 1.73 m^2^ at baseline and discovered that in persons without kidney disease at baseline, those with depressive symptoms were more likely to show signs of rapid kidney function decline over a median follow-up of 4 years and suffered 39% greater chance of rapid kidney decline, independent of other major demographic, clinical, or psychosocial covariates [7]. The second study highlighted the synergistic impact of depression in persons in the United States (US) who had already developed CKD. The 2022 retrospective analysis of 24,412 participants within the National Health and Nutrition Examination Survey 2005-2014 had a mean follow-up of 5.8 years. Persons with CKD with comorbid depression exhibited over three and a half times the hazard ratio for all-cause and cardiovascular-related mortality, followed by persons with depression without CKD, then persons with CKD only [8]. However, neither study examined the potential role of nephrotoxic medications, nor did they evaluate these relationships in the context of HIV serostatus [9].

Nephrotoxic medications, including antidepressants and antipsychotics, are commonly prescribed to manage depression in PLWH, but some of these medications have known nephrotoxic properties that may exacerbate kidney dysfunction [10]. Nephrotoxic medications may play a mediating role in the relationship between HIV and CKD, either through their nephrotoxic effects or by influencing depression-related pathways that affect kidney function. As PLWH suffer increased rates of depression compared to PLWoH, and depression has been repeatedly associated with increased morbidity and mortality in persons with CKD [11, 12], evaluating the impact of depression and nephrotoxic medication usage on kidney function in PLWH is critical.

In this study, we aimed to evaluate the prevalence and incidence of CKD among PLWH compared to people living without HIV (PLWoH), and to examine whether depression and the use of nephrotoxic medications were independently associated with CKD. We leveraged data from the Multicenter AIDS Cohort Study (MACS)/Women’s Interagency HIV Study (WIHS) Combined Cohort Study (MWCCS), the largest and longest-running prospective cohort of individuals with and without HIV in the United States, to address these questions and inform targeted clinical strategies for kidney health in this population.

Given the high prevalence of depression among PLWH, its known associations with chronic illness outcomes, and the widespread use of nephrotoxic medications, many of which have nephrotoxic potential, it is critical to examine how these factors may contribute to CKD risk in this population. We hypothesized that depression would be associated with increased CKD risk among PLWH. Additionally, we conducted causal mediation analyses to determine whether depressive symptoms and the use of nephrotoxic medications served as mediators in the relationship between HIV serostatus and CKD, thereby identifying potentially modifiable factors along this pathway.

## Materials and methods

### Study Design and Population

The MWCCS is a prospective observational cohort study designed to study the impact of chronic health conditions that affect PLWH in the US. Details have been previously described [13]. For this longitudinal analysis, baseline data were provided by the MWCCS Data Coordinating Center on participants aged 30 and older at the 13 MWCCS sites. Data were derived from three consecutive visits, identified as baseline (Visit 101), follow-up 1 (Visit 102), and follow-up 2 (Visit 103), occurring between November 16, 2020, and March 30, 2023. Specifically, Visit 101 spanned from November 16, 2020, to September 30, 2021; Visit 102 occurred from October 4, 2021, to September 30, 2022; and Visit 103 extended from October 1, 2022, to March 30, 2023. At Visit 101, 2,530 participants were enrolled as new participants. At Visit 102, 1,030 new participants joined, while 1,958 participants from Visit 101 were retained. By Visit 103, 460 new participants enrolled, and 892 participants from Visit 102 continued their participation. Across all three visits, a total of 6,870 participant records were analyzed, including 4,020 new participants and 2,850 returning participants.

This study analyzed baseline demographic, clinical, and behavioral characteristics of participants stratified by HIV serostatus to evaluate the prevalence of CKD. The analysis was conducted from 1,622 PLWH and 908 PLWoH in MWCCS. Variables were categorized and constructed based on participant demographics, socioeconomic factors, clinical diagnoses, behavioral data, and laboratory measurements. The preliminary cross-sectional analyses utilized baseline data from study subjects at visit 101, conducted between October 2020 and September 2021, across all participating MWCCS sites. Demographic data include age, gender, race, and laboratory measures for eGFR calculated using the Chronic Kidney Disease Epidemiology Collaboration (CKD-EPI) equation. Additional covariates at baseline include HIV serostatus, diabetes mellitus, hypertension, cardiovascular disease, body mass index (BMI), and substance use (alcohol, tobacco, injection drug use (IDU), cocaine, opiates). Socioeconomic factors, including education, marital status, and annual household income, were also considered. Kidney disease was indexed based on self-reported history of kidney disease diagnosis by a healthcare provider (e.g., doctor, nurse).

### Assessments of Kidney Function, Depressive Symptoms, and Nephrotoxic Medication Use

CKD was defined as eGFR <60 ml/min per 1.73 m2 (using the CKD-EPI equation) for >3 months [14], consistent with the Kidney Disease: Improving Global Outcomes (KDIGO) 2012 clinical practice guidelines for the evaluation and management of CKD. In addition, kidney function was categorized based on estimated glomerular filtration rate (eGFR) into six stages to reflect progressive kidney damage and loss of function. The stages include Kidney Damage with Normal Kidney Function (eGFR ≥ 90 ml/min/1.73 m²), Kidney Damage with Mild Loss of Kidney Function (eGFR ≥ 60 and < 90 ml/min/1.73 m²), Mild to Moderate Loss of Kidney Function (eGFR ≥ 45 and < 60 ml/min/1.73 m²), Moderate to Severe Loss of Kidney Function (eGFR ≥ 30 and < 45 ml/min/1.73 m²), Severe Loss of Kidney Function (eGFR ≥ 15 and < 30 ml/min/1.73 m²), and Kidney Failure (eGFR < 15 ml/min/1.73 m²).

Depression was measured using the Center for Epidemiologic Studies Depression (CES-D) 20-item questionnaire [15]. A cut-off score of ≥16 was used to indicate elevated depressive symptoms, consistent with established screening thresholds applied in both men and women in large cohort studies, including the MACS and WIHS cohorts [16, 17].

Nephrotoxic medications were defined as medications with established nephrotoxic properties, based on expert review and retained for analysis in this study. These medications were classified based on their potential nephrotoxic effects and nephrotoxic properties, ensuring clinical relevance in assessing exposure. The medications were assessed at the time of the visit, capturing both current use and historical medication exposure for those who may have used them in the past [18]. Medications with nephrotoxic potential were identified using a modified Delphi method, which involved expert consensus across multiple rounds to systematically review and classify medications based on their likelihood of causing kidney toxicity [19]. (Full category list is provided in Supplement Table 1). This list reflects only medications with nephrotoxic properties relevant to CKD risk.

### Assessment of Covariates

Demographics include self-reported sex at birth, race/ethnicity, and age. Race/ethnicity categories included non-Hispanic white, non-Hispanic Black, non-Hispanic other, and Hispanic. As for socioeconomic indicators, education level (highest level completed) was categorized into: less than high school, completed high school, some college, completed a four-year college degree, and completed graduate school. Marital status was classified into legally/common-law married, living with a partner, widowed, divorced, separated, never married, and other. Employment status included categories of full-time, part-time, not employed, retired, student, and disability. Income was categorized into predefined annual household income brackets. Diabetes and hypertension status were binary indicators derived from medical diagnoses or medication use. Other clinical factors included BMI, categorized into standard weight categories (e.g., underweight, normal weight, overweight, obese). Smoking status was defined as currently smoking (yes/no). Alcohol use was categorized based on binge drinking episodes since the last visit. Drug use included self-reported use of marijuana, cocaine, heroin, or other illicit drugs, as well as injection drug use.

### Statistical Analyses

Descriptive statistics were utilized to summarize baseline demographic, clinical, and behavioral characteristics of the participants stratified by HIV serostatus. For continuous variables, means and standard deviations (SD) were calculated, along with medians and ranges. Comparisons between PLWH and PLWoH were assessed using analysis of variance (ANOVA) for continuous variables. For categorical variables, frequencies and percentages were reported, and chi-square tests were employed to evaluate differences between groups.

For the prevalence analysis, we used data from Visit 101 (baseline) only. CKD prevalence was calculated as the proportion of participants meeting the CKD definition (eGFR <60 mL/min/1.73 m²) within each subgroup of interest. Results were stratified by HIV serostatus (overall, PLWH, and PLWoH) and by demographic, clinical, and behavioral characteristics.

To examine associations between risk factors and CKD, generalized estimating equations (GEE) with a Poisson distribution and log link were used to estimate relative risks while accounting for within-subject correlation across multiple study visits. Separate univariate models were fitted for each covariate to avoid potential overadjustment and misinterpretation due to collider or mediator bias (i.e., Table 2 fallacy [20]). The analysis included both categorical and continuous covariates, and relative risks (RR) and 95% confidence intervals (CI) were computed for each predictor.

**Table 1:**
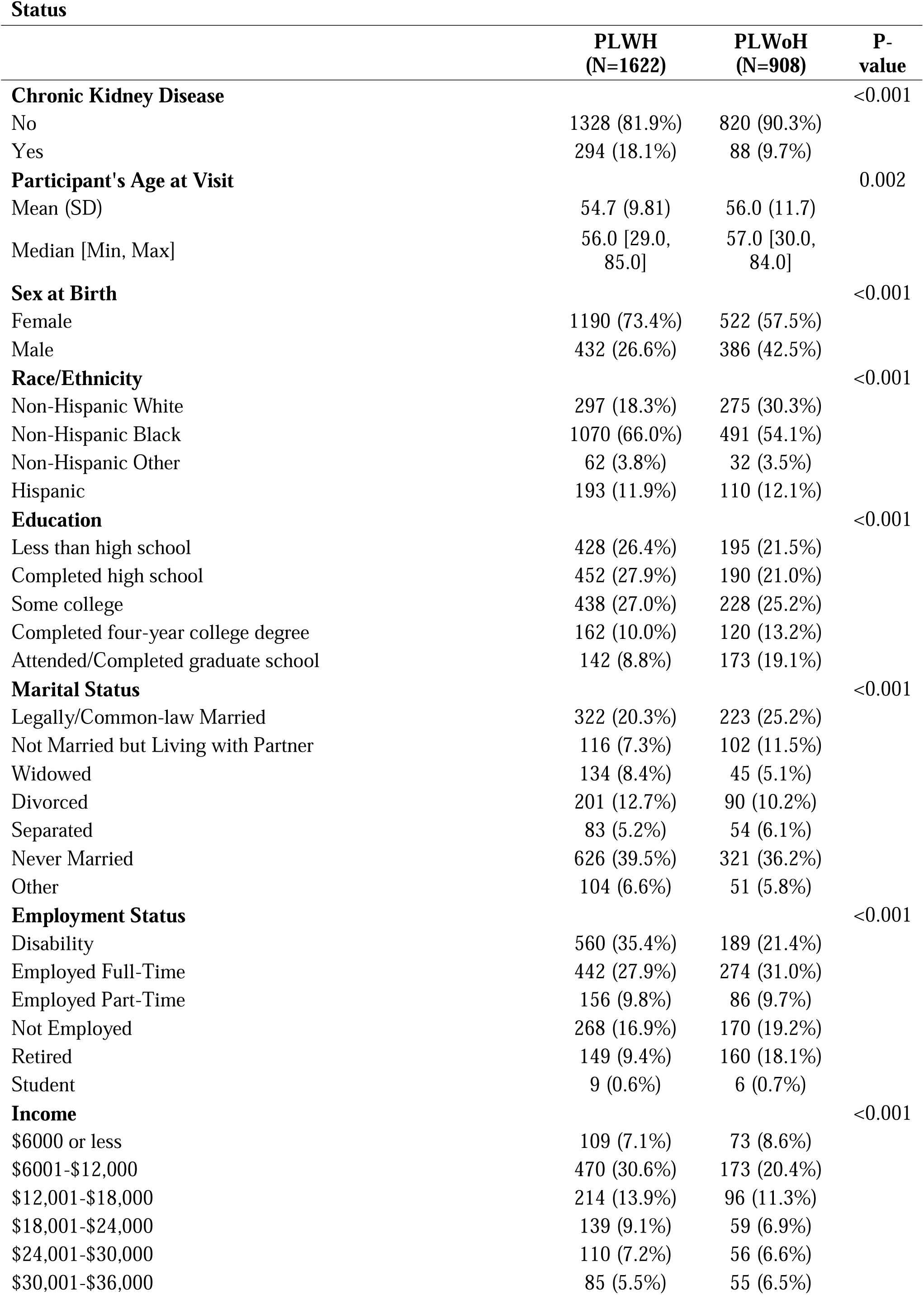
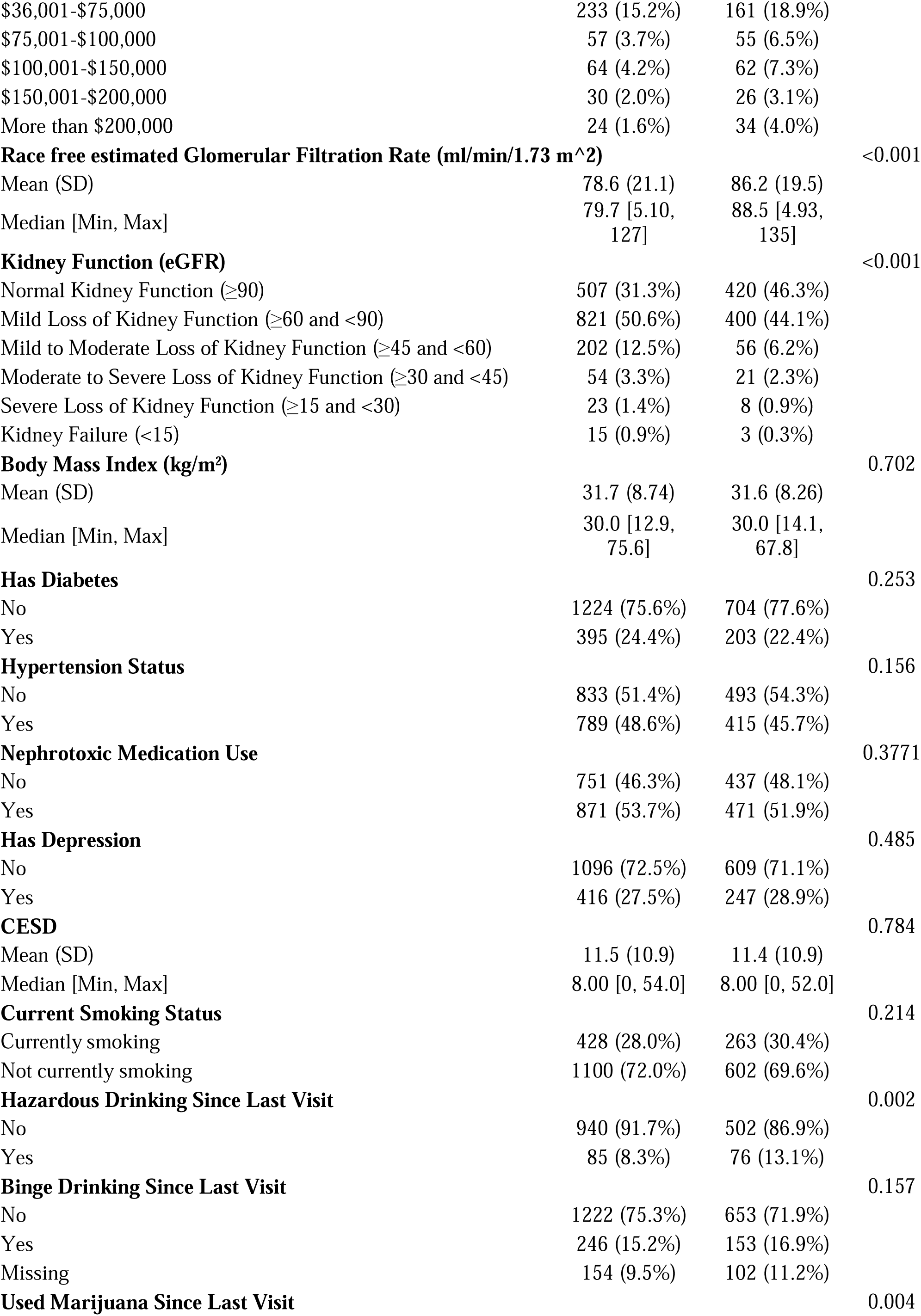
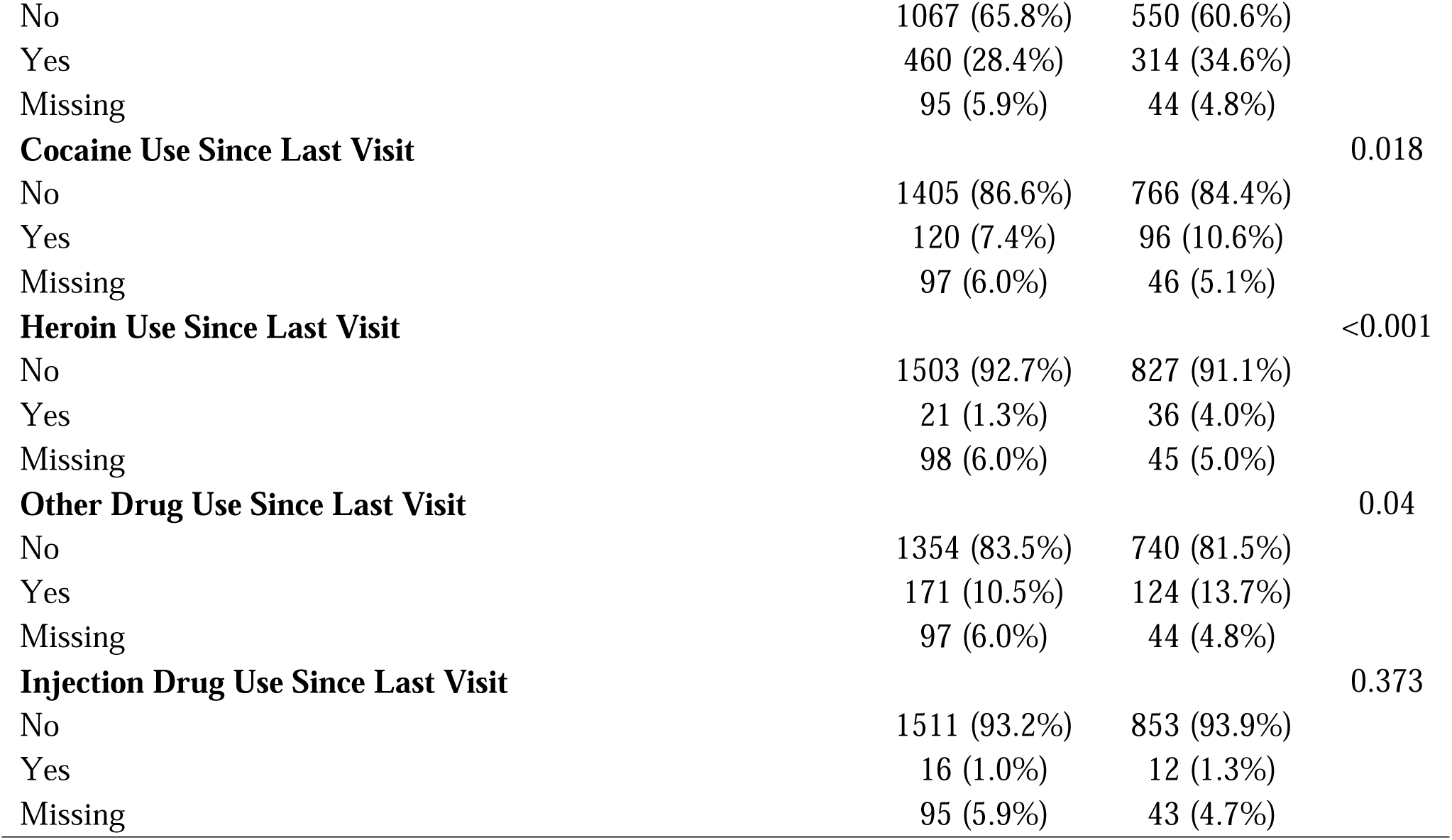
Baseline Demographic, Clinical, and Behavioral Characteristics of Participants by HIV.

**Table 2:** Prevalence of Chronic Kidney Disease by Participant Characteristics, Overall and Stratified by HIV Status.

|  |  | **Overall** |  | **PLWH** |  | **PLWoH** |  |
| --- | --- | --- | --- | --- | --- | --- | --- |
|  |  | **N** | **Prevalence (%)** | **N** | **Prevalence (%)** | **N** | **Prevalence (%)** |
| **HIV serostatus at time of interview** | HIV seronegative | 88/908 | 9.7 | - | - | - | - |
|  | HIV positive | 294/1622 | 18.1 | - | - | - | - |
| **Sex at Birth** | Female | 285/1712 | 16.7 | 224/1190 | 18.8 | 61/522 | 11.7 |
|  | Male | 97/818 | 11.9 | 70/432 | 16.2 | 27/386 | 7.0 |
| **Race/Ethnicity** | Non-Hispanic White | 70/572 | 12.2 | 51/297 | 17.2 | 19/275 | 6.9 |
|  | Non-Hispanic Black | 260/1561 | 16.7 | 201/1070 | 18.8 | 59/491 | 12.0 |
|  | Non-Hispanic Other | 17/94 | 18.1 | 15/62 | 24.2 | 2/32 | 6.3 |
|  | Hispanic | 35/303 | 11.6 | 27/193 | 14.0 | 8/110 | 7.3 |
| **Education** | Less than high school | 92/623 | 14.8 | 67/428 | 15.7 | 25/195 | 12.8 |
|  | Completed high school | 101/642 | 15.7 | 82/452 | 18.1 | 19/190 | 10.0 |
|  | Some college | 118/666 | 17.7 | 93/438 | 21.2 | 25/228 | 11.0 |
|  | Completed four-year college degree | 40/282 | 14.2 | 30/162 | 18.5 | 10/120 | 8.3 |
|  | Attended/Completed graduate school | 31/315 | 9.8 | 22/142 | 15.5 | 9/173 | 5.2 |
| **Marital Status** | Legally/Common-law Married | 73/545 | 13.4 | 49/322 | 15.2 | 24/223 | 10.8 |
|  | Not Married but Living with Partner | 24/218 | 11.0 | 18/116 | 15.5 | 6/102 | 5.9 |
|  | Widowed | 50/179 | 27.9 | 41/134 | 30.6 | 9/45 | 20.0 |
|  | Divorced | 50/291 | 17.2 | 42/201 | 20.9 | 8/90 | 8.9 |
|  | Separated | 14/137 | 10.2 | 7/83 | 8.4 | 7/54 | 13.0 |
|  | Never Married | 129/947 | 13.6 | 102/626 | 16.3 | 27/321 | 8.4 |
|  | Other | 33/155 | 21.3 | 27/104 | 26.0 | 6/51 | 11.8 |
| **Employment Status** | Not Employed | 55/438 | 12.6 | 43/268 | 16.0 | 12/170 | 7.1 |
|  | Employed Full-Time | 57/716 | 8.0 | 45/442 | 10.2 | 12/274 | 4.4 |
|  | Employed Part-Time | 26/242 | 10.7 | 21/156 | 13.5 | 5/86 | 5.8 |
|  | Retired | 71/309 | 23.0 | 47/149 | 31.5 | 24/160 | 15.0 |
|  | Student | 0/15 | 0.0 | 0/9 | 0.0 | 0/6 | 0.0 |
|  | Disability | 163/749 | 21.8 | 129/560 | 23.0 | 34/189 | 18.0 |
| **Income** | \$6000 or less | 26/182 | 14.3 | 16/109 | 14.7 | 10/73 | 13.7 |
| | \$6001-\$12,000 | 136/643 | 21.2 | 106/470 | 22.6 | 30/173 | 17.3 |
| | \$12,001-\$18,000 | 60/310 | 19.4 | 50/214 | 23.4 | 10/96 | 10.4 |
| | \$18,001-\$24,000 | 30/198 | 15.2 | 25/139 | 18.0 | 5/59 | 8.5 |
| | \$24,001-\$30,000 | 23/166 | 13.9 | 19/110 | 17.3 | 4/56 | 7.1 |
| | \$30,001-\$36,000 | 15/140 | 10.7 | 12/85 | 14.1 | 3/55 | 5.5 |
| | \$36,001-\$75,000 | 44/394 | 11.2 | 29/233 | 12.5 | 15/161 | 9.3 |
| | \$75,001-\$100,000 | 7/112 | 6.3 | 5/57 | 8.8 | 2/55 | 3.6 |
| | \$100,001-\$150,000 | 10/126 | 7.9 | 10/64 | 15.6 | 0/62 | 0.0 |
| | \$150,001-\$200,000 | 7/56 | 12.5 | 4/30 | 13.3 | 3/26 | 11.5 |
| | More than \$200,000 | 2/58 | 3.5 | 1/24 | 4.2 | 1/34 | 2.9 |
| **Has Diabetes** | No | 240/1928 | 12.5 | 190/1224 | 15.5 | 50/704 | 7.1 |
|  | Yes | 142/598 | 23.8 | 104/395 | 26.3 | 38/203 | 18.7 |
| **Hypertension Status** | No | 126/1326 | 9.5 | 107/833 | 12.9 | 19/493 | 3.9 |
|  | Yes | 256/1204 | 21.3 | 187/789 | 23.7 | 69/415 | 16.6 |
| **Has Depression** | No | 261/1705 | 15.3 | 200/1096 | 18.3 | 61/609 | 10.0 |
|  | Yes | 92/663 | 13.9 | 71/416 | 17.1 | 21/247 | 8.5 |
| **Nephrotoxic Medication Use** | No | 124/1188 | 10.4 | 99/751 | 13.2 | 25/437 | 5.7 |
|  | Yes | 258/1342 | 19.2 | 195/871 | 22.4 | 63/471 | 13.4 |
| **Current Smoking Status** | Not currently smoking | 248/1702 | 14.6 | 198/1100 | 18.0 | 50/602 | 8.3 |
|  | Currently smoking | 105/691 | 15.2 | 72/428 | 16.8 | 33/263 | 12.6 |
| **Binge Drinking Since Last Visit** | No | 294/1875 | 15.7 | 229/1222 | 18.7 | 65/653 | 10.0 |
|  | Yes | 38/399 | 9.5 | 29/246 | 11.8 | 9/153 | 5.9 |
| **Used Marijuana Since Last Visit** | No | 261/1617 | 16.1 | 206/1067 | 19.3 | 55/550 | 10.0 |
|  | Yes | 92/774 | 11.9 | 64/460 | 13.9 | 28/314 | 8.9 |
| **Cocaine Use Since Last Visit** | No | 315/2171 | 14.5 | 247/1405 | 17.6 | 68/766 | 8.9 |
|  | Yes | 36/216 | 16.7 | 22/120 | 18.3 | 14/96 | 14.6 |
| **Heroin Use Since Last Visit** | No | 343/2330 | 14.7 | 266/1503 | 17.7 | 77/827 | 9.3 |
|  | Yes | 9/57 | 15.8 | 4/21 | 19.1 | 5/36 | 13.9 |
| **Other Drug Use Since Last Visit** | No | 323/2094 | 15.4 | 247/1354 | 18.2 | 76/740 | 10.3 |
|  | Yes | 29/295 | 9.8 | 22/171 | 12.9 | 7/124 | 5.7 |
| **Injection Drug Use Since Last Visit** | No | 349/2364 | 14.8 | 268/1511 | 17.7 | 81/853 | 9.5 |
|  | Yes | 4/28 | 14.3 | 2/16 | 12.5 | 2/12 | 16.7 |

The GEE models incorporated an exchangeable working correlation matrix and accounted for repeated measurements clustered within individuals. Variables examined included HIV serostatus (PLWH vs. PLWoH), age, sex, race/ethnicity, marital status, education level, income, diabetes, hypertension, depression, nephrotoxic medication use, smoking, alcohol use, other substance use, and BMI. For categorical variables with more than two levels (e.g., education, marital status, race/ethnicity), comparisons were made relative to the reference group. Statistical significance was determined using Wald tests, and model fit was evaluated using empirical standard errors and robust variance estimates.

In addition, we performed counterfactual mediation analysis to quantify the pathway-specific effects of HIV on CKD through depression and nephrotoxic medication use. Using a binomial model with a logit link, we estimated the *total effect (TE)*, the *natural direct effect (NDE)*, and the *natural indirect effect (NIE)* on the odds-ratio scale, adjusting for sex, race, marital status, diabetes, hypertension, smoking, and substance use. The NDE represents the effect of HIV on CKD if the mediator were set to the level it would naturally take under the reference (PLWoH) exposure; the NIE represents the portion operating through the mediator. We allowed for exposure mediator interaction. The primary goal was to determine the extent to which depression or nephrotoxic medication use mediated the pathway from HIV to CKD, while accounting for potential interaction between the exposure and the mediator. Bootstrap resampling with 1,000 iterations was used to derive robust 95% confidence intervals. A two-sided p-value of <0.05 was considered statistically significant.

## Results

### Study Participants and Baseline Characteristics

The study included 1,622 PLWH and 908 PLWoH participants (Table 1). PLWH had a slightly younger mean age (54.7 years) compared to PLWoH (56.0 years, p=0.002). A higher proportion of PLWH were female (73.4%) compared to PLWoH (57.5%, p<0.001). The majority of participants were non-Hispanic Black individuals (66.0% of PLWH and 54.1% of PLWoH). Socioeconomic indicators revealed disparities in education and income levels, with PLWH more likely to have lower educational attainment and income levels than PLWoH (p<0.001).

Significant differences were observed in the prevalence of CKD, with 18.1% of PLWH affected compared to 9.7% of PLWoH participants (p<0.001). Additionally, measures of kidney function indicated worse outcomes among the PLWH, with fewer participants retaining normal kidney function (31.3% vs. 46.3%, p<0.001).

### Prevalence of CKD overall and stratified by HIV serostatus

Table 2 presents CKD prevalence across demographic, clinical, and behavioral characteristics, stratified by HIV serostatus. Higher CKD prevalence was observed among females, individuals with lower educational attainment, and persons identifying as non-Hispanic Black or non-Hispanic Other. Among marital categories, widowed and “other” individuals exhibited elevated CKD rates. Participants who were retired or had a disability showed higher CKD prevalence compared to those employed full-time. Behavioral factors such as current smoking, binge drinking, and illicit drug use were associated with higher CKD prevalence, particularly among PLWH.

Depression showed a nuanced association with CKD. Among individuals without depression, CKD prevalence was 15.3%, higher in PLWH (18.3%) than PLWoH (10%). Among those with depression, overall CKD prevalence was 13.9%, also higher in PLWH (17.1%) than PLWoH (8.5%). At Visit 101, 1,342 participants (53%) were classified as using nephrotoxic medications (Table 3). Nephrotoxic medication use was 51.9% in PLWoH and 53.7% in PLWH.

**Table 3:** Prevalence of Nephrotoxic Medication Use by Visit and HIV Status.

| **Visit Number** | **HIV Status** | **Total**<br>**Participants** | **Participants**<br>**Using**<br>**Nephrotoxic**<br>**Medications (n)** | **Nephrotoxic**<br>**Medication Use**<br>**(%)** |
| --- | --- | --- | --- | --- |
| 101 | Overall | 2,530 | 1,342 | 53.0 |
| 101 | HIV- | 908 | 471 | 51.9 |
| 101 | HIV+ | 1,622 | 871 | 53.7 |
| 102 | Overall | 2,985 | 1,624 | 54.4 |
| 102 | HIV- | 1,074 | 566 | 52.7 |
| 102 | HIV+ | 1,911 | 1,058 | 55.4 |
| 103 | Overall | 1,351 | 699 | 51.7 |
| 103 | HIV- | 440 | 213 | 48.4 |
| 103 | HIV+ | 911 | 486 | 53.4 |

\*Nephrotoxic medication use was defined based on expert review of medications with established nephrotoxic properties (see Supplement Table 1).

### Generalized Estimating Equations (GEE) Analysis of Kidney Function Decline

Table 4 presents the relationship between HIV status, depression, and chronic kidney disease (CKD). PLWH had a significantly higher risk of kidney function decline compared to PLWoH (RR□=□1.37, 95% CI: 1.28- 1.48, p□<□0.001). Age was also positively associated with poorer kidney function, with each additional year of age associated with a 3% increase in CKD risk (RR□=□1.03 per year, 95% CI: 1.03-1.03, p□<□0.001).

**Table 4.** GEE Analysis Results: Factors Associated with Chronic Kidney Disease.

| **Variable** | **RR** | **95% CI** | **P-value** |
| --- | --- | --- | --- |
| HIV Serostatus (HIV Positive vs. HIV Negative) | **1.37** | **(1.48, 1.28)** | **<.0001** |
| Age (per unit increase) | **1.03** | **(1.03, 1.03)** | **<.0001** |
| Sex (Female vs. Male) | **1.16** | **(1.23, 1.08)** | **<.0001** |
| Race: Non-Hispanic Black (vs. Non-Hispanic White) | **1.19** | **(1.11, 1.27)** | **<.0001** |
| Race: Hispanic (vs. Non-Hispanic White) | **2.03** | **(1.86, 2.21)** | **<.0001** |
| Race: Non-Hispanic Other (vs. Non-Hispanic White) | **2.64** | **(2.31, 3.02)** | **<.0001** |
| Education: Completed high school (HS) vs. < HS | 0.83 | (0.68, 1.03) | 0.087 |
| Education: Some college vs. < HS | 0.87 | (0.72, 1.07) | 0.184 |
| Education: Completed/graduate college vs. < HS | 0.81 | (0.64, 1.02) | 0.069 |
| Education: Attended/completed post-grad vs. < HS | 0.86 | (0.69, 1.06) | 0.163 |
| Marital: Not Married but Living with Partner (vs. Married) | 1.16 | (1.01, 1.32) | **0.0364** |
| Marital: Widowed (vs. Married) | 1.16 | (0.97, 1.4) | 0.1059 |
| Marital: Divorced (vs. Married) | 1.07 | (0.93, 1.25) | 0.3432 |
| Marital: Separated (vs. Married) | 1.16 | (1.0, 1.36) | 0.0556 |
| Marital: Never Married (vs. Married) | 1.01 | (0.9, 1.12) | 0.8762 |
| Marital: Other (vs. Married) | 1.02 | (0.89, 1.18) | 0.7433 |
| Income (per unit increase) | 0.99 | (0.98, 1.0) | **0.005** |
| Diabetes (Yes vs. No) | 1.26 | (1.17, 1.35) | **<.0001** |
| Hypertension (Yes vs. No) | 1.06 | (0.97, 1.15) | 0.2043 |
| Depression (Yes vs. No) | 1.04 | (0.96, 1.14) | 0.3355 |
| Nephrotoxic Medication (Yes vs. No) | 1.04 | (0.96, 1.14) | 0.3355 |
| Current Smoker (vs. Non-Smoker) | 1.09 | (1.31, 0.91) | 0.3352 |
| Marijuana Use (Yes vs. No) | 1.1 | (0.96, 1.25) | 0.1655 |
| Cocaine Use (Yes vs. No) | 0.92 | (0.75, 1.12) | 0.3918 |
| Heroin Use (Yes vs. No) | 0.69 | (0.28, 1.71) | 0.4257 |
| Other Drug Use (Yes vs. No) | 0.96 | (0.76, 1.21) | 0.7075 |
| Injection Drug Use (Yes vs. No) | 1 | (0.97, 1.03) | 0.9276 |
| BMI (per unit increase) | 1.02 | (0.99, 1.05) | 0.2426 |

Among racial/ethnic groups, non-Hispanic Black individuals had a significantly higher risk (RR□=□1.19, 95% CI: 1.11-1.27, p□<□0.001), and Hispanic individuals had more than twice the risk compared to non-Hispanic White individuals (RR□=□2.03, 95% CI: 1.86-2.21, p□<□0.001). Additionally, non-Hispanic Other individuals also showed an elevated risk (RR□=□2.64, 95% CI: 2.31-3.02, p□<□0.001). Sex was also significantly associated, with females having higher CKD risk compared to males (RR□=□1.16, 95% CI: 1.08-1.23, p□<□0.001). Higher income was associated with lower CKD risk (RR□=□0.99, 95% CI: 0.98-1.00, p□=□0.005). Among chronic conditions, diabetes was significantly associated with increased risk of CKD (RR□=□1.26, 95% CI: 1.17-1.35, p□<□0.001), while hypertension was not statistically significant (RR□=□1.06, 95% CI: 0.97-1.15, p□=□0.204).

Depression or Nephrotoxic medication use mediates the relationship between HIV Status and CKD. We conducted a mediation analysis to evaluate whether depression and nephrotoxic medication use mediated the relationship between HIV status and CKD. The total OR of CKD associated with HIV status was 2.28 (95% CI: 1.88-2.77, p□<□0.0001; Fig 1), indicating a strong association between HIV and CKD. The natural direct effect (NDE), i.e., portion of the HIV–CKD association not operating through nephrotoxic medication use, was similar in magnitude (OR□=□2.25, 95% CI: 1.84-2.73, p < 0.0001).

**Figure 1.**
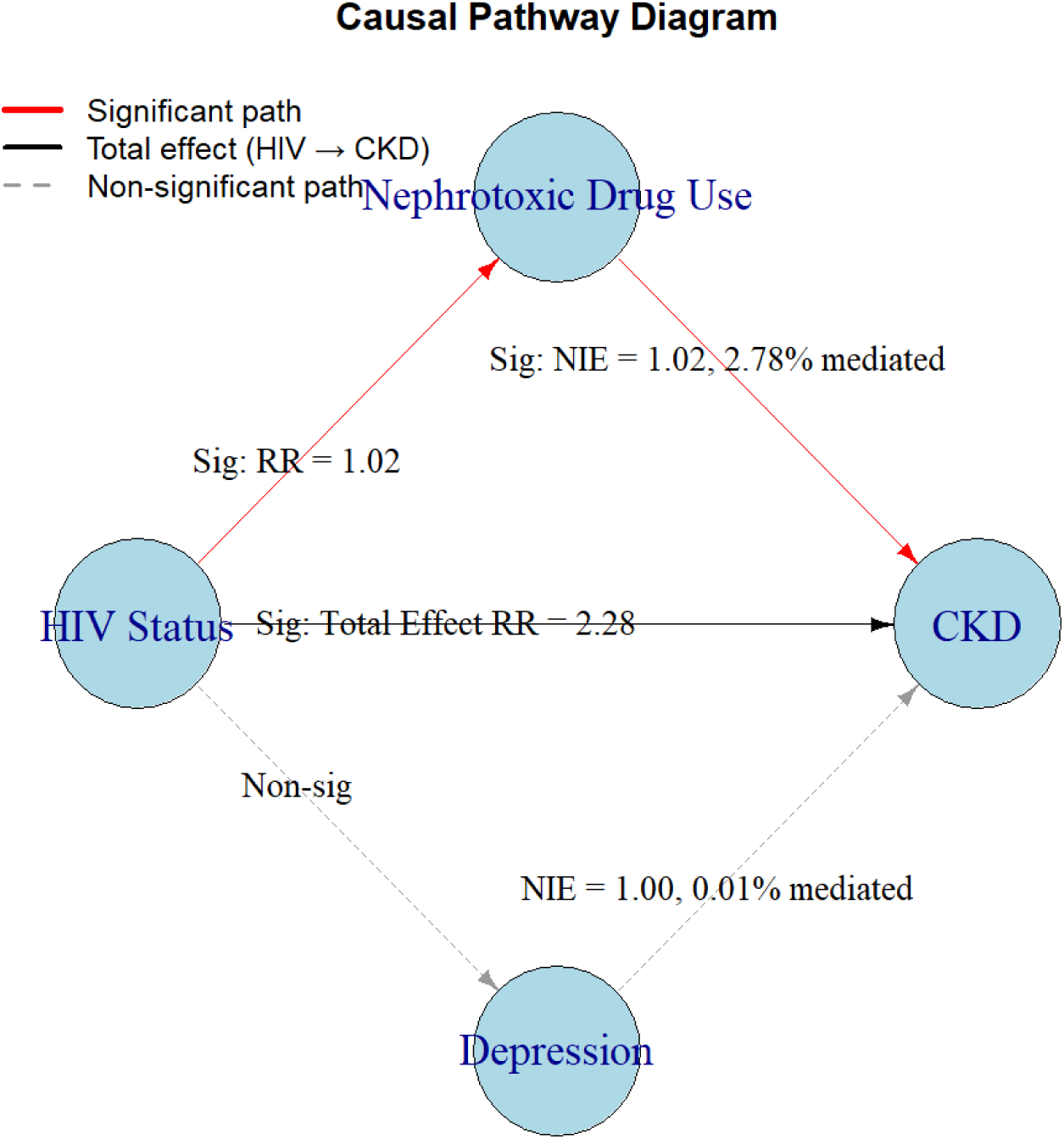
Causal Pathway Diagram.

The natural indirect effect (NIE) through nephrotoxic drug use was modest but statistically significant (OR = 1.02, 95% CI: 1.00-1.03, p = 0.019), accounting for ∼2.78% of the total effect. Most of the association was explained by the direct pathway, with a small yet meaningful portion mediated through nephrotoxic drug exposure. Importantly, the interaction between HIV and nephrotoxic drug use was statistically significant (17.83%, p = 0.0002), suggesting that nephrotoxic drug exposure further increased CKD risk among people living with HIV.

## Discussion

While previous studies have suggested a strong link between depression and CKD progression, our analysis did not find a direct relationship. This lack of association may be due to effective treatment and management of depression in this cohort, which could have mitigated the effects on kidney function. In contrast, our mediation analysis revealed that nephrotoxic medication use played a modest but significant role in mediating the relationship between HIV and CKD. These agents have well-documented nephrotoxic effects that could exacerbate kidney damage. Thus, the mediating role of nephrotoxic medications in the HIV - CKD pathway is likely driven by their nephrotoxicity. Importantly, the interaction between HIV and nephrotoxic drug use was statistically significant, suggesting that nephrotoxic drug exposure amplifies CKD risk in PLWH. These findings highlight the need for clinicians to routinely assess kidney function when prescribing or managing nephrotoxic medications in PLWH and to weigh renal risks alongside clinical benefits as part of personalized HIV care.

Following this key finding, we examined the broader relationships between HIV serostatus, depression, nephrotoxic medication use, and CKD. First, consistent with prior research [1], we observed a high prevalence of CKD, and a significantly higher prevalence of CKD in PLWH (18.1%) compared to PLWoH (9.7%). This disparity remained significant after adjusting for demographic, clinical, and behavioral covariates. Furthermore, the longitudinal analysis using GEE confirmed that HIV serostatus was a strong predictor of kidney function decline. Our findings are congruent with prior studies documenting that kidney disease in PLWH is a common and serious complication, with reports documenting kidney disease as the fourth leading cause of death in patients with AIDS prior to the use of effective ART and among individuals not using ART. Although HIV-associated nephropathy (HIVAN), especially in individuals carrying APOL1 risk alleles, was once the most common form of kidney injury in PLWH, its incidence has declined dramatically in the current ART era [21]. Today, other forms of CKD, such as diabetic kidney disease and hypertensive nephrosclerosis, are more frequently observed [14, 21–25]. HIV can also lead to renal injury through mechanisms such as HIV-associated thrombotic microangiopathy (TMA), acute interstitial nephritis, immune complex-mediated glomerular disease, and sepsis-related acute tubular necrosis (ATN) due to immunosuppression [26].

Fortunately, compared to people without HIV, kidney dysfunction in PLWH has declined with the use of modern ART and may now progress at a similar or slower rate compared to those without HIV. However, many medications used to treat HIV and the complications of HIV can result in renal injury. Protease inhibitors, such as atazanavir, have been associated with crystal-induced obstructive acute kidney injury (AKI) [27]. Tenofovir (a nucleoside reverse transcriptase inhibitors) use has been implicated in tubulopathies causing Fanconi syndrome and nephrogenic diabetes insipidus. Moreover, the presence of traditional risk factors such as hypertension, diabetes, and older age, remained critical determinants of CKD progression across both groups in our study.

Of note, individuals in our cohort with depression had a slightly lower prevalence of CKD compared to those without depression among PLWH. However, among PLWoH, CKD prevalence was also lower in those with depression compared to those without. These findings contrast with prior literature that suggests a strong link between depression and CKD progression [28–31], that is, there is a higher prevalence of depression as CKD worsens. One possible explanation for this discrepancy is that the MWCCS cohort includes individuals with more stable access to care and effective management of both HIV and mental health conditions. This may have attenuated the impact of depression on CKD progression. Studies such as those by Capuron et al. and Lustman et al. have suggested that while somatic depressive symptoms (e.g., fatigue, appetite loss) are more prevalent in conditions like diabetes, cognitive symptoms (e.g., sadness, hopelessness) may be more prominent in populations of persons with [12, 32]. In contrast, another group showed that in persons with cancer, the most optimal diagnostic tool for the identification of depression contained *both* somatic and cognitive symptoms (late insomnia, agitation, psychic anxiety, diurnal mood variation, depressed mood, and decreased libido) of the Hamilton Depression Rating Scale [33]. However, the lack of a significant mediating effect of depression on CKD progression in our analysis suggests that, although depression is highly prevalent among individuals with CKD, it may not independently contribute to disease progression. One possible explanation is that participants with elevated depressive symptoms may have benefited from better access to supportive services or clinical follow-up that attenuated CKD risk. Nevertheless, because depression in this study was defined solely based on CES-D symptom scores rather than clinical diagnosis or treatment history, we cannot determine whether participants were receiving appropriate or effective depression management. The potential modifying effect of depression on CKD risk, particularly among individuals with HIV, remains unclear and warrants further investigation in future studies that incorporate both symptom-based and treatment-based assessments.

Despite the strong association between HIV and CKD, the mediation analysis indicated that nephrotoxic medication use accounted for a small but significant proportion of this relationship. These findings align with prior studies suggesting that certain nephrotoxic drugs, including commonly used antiretrovirals and other agents, may contribute to kidney injury over time [30, 34–37].

### Strengths and Limitations

Our study has several strengths. A key strength is our use of mediation analysis to disentangle the direct and indirect effects of HIV on CKD, thereby allowing us to gain new insights into the role of nephrotoxic medication exposure in this association among PLWH. Additionally, the longitudinal design allowed us to assess CKD progression over time and explore differences in HIV serostatus, depression, and medication use. The MWCCS cohort also represents a deeply phenotyped, diverse population.

However, there are limitations to consider. The observational nature of our study precludes causal inferences, and unmeasured confounding variables, such as levels of inflammation markers (e.g., C-reactive protein) and alternative kidney function measures (e.g., cystatin C), were not available in our dataset. Inflammation, medication adherence, and healthcare access may influence both depression and CKD risk. These markers may have provided a more accurate estimation of kidney function, particularly in individuals with low muscle mass [38]. Medication data were limited to nephrotoxic drug exposures captured in the MWCCS cohort. As a result, while we were able to quantify the mediating effect of nephrotoxic medications overall, we could not evaluate the impact of non-nephrotoxic psychiatric medications because they were not captured in the coded data during the study window. This restricts the generalizability of our findings and should be interpreted accordingly.

## Conclusion

In conclusion, our study reaffirms the strong association between HIV and CKD, even among PLWH successfully treated with ART, with most of the risk explained by direct effects rather than mediation through depression. While depression was highly prevalent among persons with CKD, it was not independently associated with CKD in multivariable models. In contrast, nephrotoxic medication use played a modest but significant mediating role, suggesting that the contribution of medications to CKD risk in PLWH is largely driven by their renal toxicity. These findings highlight the need for careful renal monitoring in patients prescribed medications with nephrotoxic potential. Future research should further explore the biological mechanisms linking HIV, depression, and kidney disease, with particular attention to both medication-related toxicity and systemic inflammatory pathways.

## Ethics Statement

This study used data from the NIH-funded MWCCS cohort. All participants provided written informed consent at each site’s institutional review board (IRB)-approved protocol.

## Data Availability

MWCCS data are available to qualified researchers through the MWCCS Data Analysis and Coordination Center (https://mwccs.org/data-access).

## Data Availability

The data underlying the results presented in this study are available from the Multicenter AIDS Cohort Study / Women’s Interagency HIV Study Combined Cohort Study (MWCCS) Data Analysis and Coordination Center (DACC) for researchers who meet the criteria for access to confidential data. Data can be requested at https://statepi.jhsph.edu/mwccs/. The MWCCS data are not publicly available because of institutional and federal privacy protections for human subjects, but de-identified datasets are accessible to qualified researchers through a formal data access process administered by the DACC.

## Acknowledgement

The authors gratefully acknowledge the contributions of the study participants and dedication of the staff at the MWCCS sites.

