## Supplemental Table 1 for "HIV, Nephrotoxic Medications, and Chronic Kidney Disease: Prevalence, Risk Factors, and Mediation Analyses Among People With and Without HIV Enrolled in the Multicenter AIDS Cohort Study (MACS) / Women’s Interagency HIV Study (WIHS) Combined Cohort Study"

| **Supplementary Table: Classification of Medications by Nephrotoxic Properties** | | |
| --- | --- | --- |
| **No.** | **Medication Name** | **Nephrotoxic Drug** |
| **1** | ASA | Yes |
| **2** | Acetazolamid | Yes |
| **3** | Acyclovir | Yes |
| **4** | Advil | Yes |
| **5** | Aleve | Yes |
| **6** | Aleve-D | Yes |
| **7** | Allopurinol | Yes |
| **8** | Alopurinol | Yes |
| **9** | Amox | Yes |
| **11** | Anacin | Yes |
| **12** | Aspirin | Yes |
| **13** | Azor | Yes |
| **14** | Bactrim | Yes |
| **15** | Bayer Aspirin | Yes |
| **16** | Bayer Back & Body | Yes |
| **17** | Benicar | Yes |
| **18** | Biktarvy | Yes |
| **19** | Bisoprolol-Hydrochlorothiazide | Yes |
| **20** | Celebrex | Yes |
| **21** | Cipro | Yes |
| **22** | Ciprofloxacine | Yes |
| **23** | Complera | Yes |
| **24** | Descovy | Yes |
| **25** | Diclophenac | Yes |
| **27** | Ecotrin | Yes |
| **28** | Entresto | Yes |
| **29** | Envarsus | Yes |
| **30** | Excedrin | Yes |
| **31** | Exforge HCT | Yes |
| **32** | Farxiga | Yes |
| **33** | Furosemid | Yes |
| **34** | Furosemide | Yes |
| **35** | Gabapentine | Yes |
| **36** | Genvoya | Yes |
| **39** | HCTZ | Yes |
| **40** | HCTZ Triamterene | Yes |
| **41** | HYDROCHLOROTHIAZIDE/LISINOPRIL | Yes |
| **42** | Hydrochlorothiazide | Yes |
| **43** | Hydroxychlorochin | Yes |
| **44** | Hyzaar | Yes |
| **45** | Ibuprofen | Yes |
| **46** | Ibuprophen | Yes |
| **48** | Lasix | Yes |
| **49** | Lisinopril | Yes |
| **50** | Losartan | Yes |
| **51** | METFORMIN | Yes |
| **52** | Metformin | Yes |
| **53** | Metformine | Yes |
| **54** | Motrin | Yes |
| **55** | Naprosin | Yes |
| **56** | Naprosyn | Yes |
| **57** | Naproxen | Yes |
| **58** | Neurontin | Yes |
| **59** | Nexplanon | Yes |
| **60** | OMEPRAZOLE | Yes |
| **61** | Omeprazol | Yes |
| **62** | Omeprazole | Yes |
| **63** | Prevacid | Yes |
| **64** | Prilosec | Yes |
| **65** | Prinivil | Yes |
| **66** | Prinzide | Yes |
| **67** | Protonix | Yes |
| **68** | Reclast | Yes |
| **69** | Reglan | Yes |
| **70** | Remdesivir | Yes |
| **71** | SPIRONOLACTONE | Yes |
| **72** | SULFAMETHOXAZOLE/TRIMETHOPRIM | Yes |
| **73** | Spirolactone | Yes |
| **74** | Sulfamethoxazole | Yes |
| **75** | Symtuza | Yes |
| **76** | Tivicay | Yes |
| **77** | Tribenzor | Yes |
| **78** | Triumeq | Yes |
| **79** | Truvada | Yes |
| **80** | Valtrex | Yes |
| **81** | Vemlidy | Yes |
| **82** | Vimpat | Yes |
| **83** | Voltaren | Yes |
| **84** | Zovirax | Yes |
| **85** | acetaZOLAMIDE | Yes |
| **86** | acetazolamide | Yes |
| **87** | acyclovir | Yes |
| **88** | acyclovir 400 MG | Yes |
| **89** | advil | Yes |
| **90** | aleve | Yes |
| **91** | allopurinol | Yes |
| **92** | amantadine | Yes |
| **93** | amoxicillin / clavulanate | Yes |
| **94** | asa | Yes |
| **95** | aspirin | Yes |
| **96** | aspirin 81 MG | Yes |
| **97** | aspirin 81 mg | Yes |
| **98** | bactrim | Yes |
| **99** | bayer aspirin | Yes |
| **100** | benazapril | Yes |
| **101** | biktarvy | Yes |
| **102** | bisoprolol/hydrochlorothiazide | Yes |
| **103** | celebrex | Yes |
| **104** | celecoxib | Yes |
| **105** | descovy | Yes |
| **106** | dicrofenac | Yes |
| **107** | digoxin | Yes |
| **108** | dihydrochlorothiazide | Yes |
| **109** | enalapril | Yes |
| **110** | entresto | Yes |
| **111** | excedrin | Yes |
| **112** | farxiga | Yes |
| **113** | fosinopril/hydrochlorothiazide | Yes |
| **114** | furosemide | Yes |
| **115** | gabapentin | Yes |
| **116** | genvoya | Yes |
| **118** | hctz | Yes |
| **119** | hydroCHLOROthiazide | Yes |
| **120** | hydroCHLOROthiazide / triamterene | Yes |
| **121** | hydrochlorothiazide | Yes |
| **122** | hydroxychloroquine | Yes |
| **123** | hyzaar | Yes |
| **124** | ibuprofen | Yes |
| **125** | ibuprofen 800 MG | Yes |
| **126** | ibuprophen | Yes |
| **127** | indomethacin | Yes |
| **129** | ketorolac | Yes |
| **130** | ketorolac tromethamine | Yes |
| **131** | lansoprazol | Yes |
| **132** | lasix | Yes |
| **133** | lisinopril | Yes |
| **134** | lithium | Yes |
| **135** | lithium carbonate | Yes |
| **136** | losartan | Yes |
| **137** | losartan potassium | Yes |
| **138** | metFORMIN | Yes |
| **139** | metformin | Yes |
| **140** | metformin hydrochloride | Yes |
| **141** | metformine | Yes |
| **142** | methotrexate | Yes |
| **143** | mobic | Yes |
| **144** | motrin | Yes |
| **145** | naprosin | Yes |
| **146** | naproxen | Yes |
| **147** | neomycin | Yes |
| **148** | neurontin | Yes |
| **149** | nexplanon | Yes |
| **150** | nitrofurantoin | Yes |
| **151** | odefsey | Yes |
| **152** | ofloxacin | Yes |
| **153** | olmesartan | Yes |
| **154** | omeprazole | Yes |
| **155** | omesartan | Yes |
| **156** | pantoprazole | Yes |
| **158** | prevacid | Yes |
| **159** | prilosec | Yes |
| **160** | prinivil | Yes |
| **161** | protonix | Yes |
| **162** | quinapril | Yes |
| **163** | ramipril | Yes |
| **164** | relafen | Yes |
| **165** | remdesivir | Yes |
| **166** | rifampin | Yes |
| **167** | sotalol | Yes |
| **168** | spirolactone | Yes |
| **169** | spironolactone | Yes |
| **170** | spirulina | Yes |
| **171** | sulfamethoxazole | Yes |
| **172** | sulfamethoxazole-trimethoprim | Yes |
| **173** | sulfamethoxazole/trimethoprim | Yes |
| **174** | sulfasalazine | Yes |
| **175** | sulindac | Yes |
| **176** | tenofovir | Yes |
| **177** | tivicay | Yes |
| **178** | tribenzor | Yes |
| **179** | truvada | Yes |
| **180** | valACYclovir | Yes |
| **181** | valaciclovir | Yes |
| **182** | valacyclovir | Yes |
| **183** | valacyclovir 500 mg | Yes |
| **184** | valacyclovir HCl | Yes |
| **185** | valsartan | Yes |
| **186** | valtrex | Yes |
| **187** | vancomycin | Yes |
| **188** | vemlidy | Yes |
| **189** | vimpat | Yes |
| **190** | viread | Yes |
| **191** | voltaren | Yes |
| **192** | zovirax | Yes |
